# Personalized Predictive Models for Symptomatic COVID-19 Patients Using Basic Preconditions: *Hospitalizations, Mortality, and the Need for an ICU or Ventilator*

**DOI:** 10.1101/2020.05.03.20089813

**Authors:** Salomón Wollenstein-Betech, Christos G. Cassandras, Ioannis Ch. Paschalidis

**Affiliations:** Division of Systems Engineering, Department of Electrical and Computer Engineering, Boston University, Boston, MA 02215; Department of Electrical and Computer Engineering, Boston University, Boston, MA 02215; Department of Biomedical Engineering, Boston University, Boston, MA 02215

**Keywords:** Predictive models, COVID-19, coronavirus, SARS-CoV-2, hospitalization, mortality, ICU, Ventilator, Electronic Health Records (EHRs)

## Abstract

**Background:** The rapid global spread of the virus SARS-CoV-2 has provoked a spike in demand for hospital care. Hospital systems across the world have been over-extended, including in Northern Italy, Ecuador, and New York City, and many other systems face similar challenges. As a result, decisions on how to best allocate very limited medical resources have come to the forefront. Specifically, under consideration are decisions on who to test, who to admit into hospitals, who to treat in an Intensive Care Unit (ICU), and who to support with a ventilator. Given today’s ability to gather, share, analyze and process data, personalized predictive models based on demographics and information regarding prior conditions can be used to (1) help decision-makers allocate limited resources, when needed, (2) advise individuals how to better protect themselves given their risk profile, (3) differentiate social distancing guidelines based on risk, and (4) prioritize vaccinations once a vaccine becomes available.

**Objective:** To develop personalized models that predict the following events: (1) hospitalization, (2) mortality, (3) need for iCu, and (4) need for a ventilator. To predict hospitalization, it is assumed that one has access to a patient’s basic preconditions, which can be easily gathered without the need to be at a hospital. For the remaining models, different versions developed include different sets of a patient’s features, with some including information on how the disease is progressing (e.g., diagnosis of pneumonia).

**Materials and Methods:** Data from a publicly available repository, updated daily, containing information from approximately 91,000 patients in Mexico were used. The data for each patient include demographics, prior medical conditions, SARS-CoV-2 test results, hospitalization, mortality and whether a patient has developed pneumonia or not. Several classification methods were applied, including robust versions of logistic regression, and support vector machines, as well as random forests and gradient boosted decision trees.

**Results:** Interpretable methods (logistic regression and support vector machines) perform just as well as more complex models in terms of accuracy and detection rates, with the additional benefit of elucidating variables on which the predictions are based. Classification accuracies reached 61%, 76%, 83%, and 84% for predicting hospitalization, mortality, need for ICU and need for a ventilator, respectively. The analysis reveals the most important preconditions for making the predictions. For the four models derived, these are: (1) for hospitalization: age, gender, chronic renal insufficiency, diabetes, immunosuppression; (2) for mortality: age, SARS-CoV-2 test status, immunosuppression and pregnancy; (3) for ICU need: development of pneumonia (if available), cardiovascular disease, asthma, and SARS-CoV-2 test status; and (4) for ventilator need: ICU and pneumonia (if available), age, gender, cardiovascular disease, obesity, pregnancy, and SARS-CoV-2 test result.

## 1 Introduction

Currently, the world is facing a health and economic crisis due to the spread of the virus SARS-CoV-2 which causes a disease referred to as COVID-19 [1]. By the end of April 2020, the virus has spread to over 3.3 million people worldwide and has killed over 230,000 [2,3]. During this pandemic, governments and hospitals have struggled to allocate scarce resources, including tests, treatment in intensive care units (ICUs) and ventilators [4,5].

As the virus continues to spread, predicting hospitalizations, mortality, and other patient outcomes becomes important for several reasons: (i) using risk profiles to inform decisions on who should be tested (for the virus and/or antibodies) and at which frequency, (ii) providing more accurate estimates of who is more likely to be hospitalized and the type of care they may need, (iii) informing plans for staffing, resources, and prioritizing the level of care in extremely resource-constrained settings. Equally importantly, as societies adapt to the pandemic, predictive models can (i) assess individual risk so that social distancing measures can transition from “blanket” to more targeted (e.g., deciding who can return to work, who is advised to stay at home, who should be tested, etc.) and (ii) direct policy decisions on who should receive priority for vaccination, which will be critical as initial vaccine production may not suffice to vaccinate everybody.

To develop predictive models, we leverage supervised machine learning methods that learn from given examples of predictive variables and associated outcomes – the so called training set. Performance is then evaluated on a separate test set. In the specific application of interest, we will focus on classification, a setting where the outcome is binary, e.g., someone is hospitalized or not.

Many models have been used to predict a patient admission to a hospital, mortality and other health care applications based on comorbidities. Some examples include: predicting morbidity of patients with chronic obstructive pulmonary disease [6], febrile neutropenia [7], as well as classifying the hospitalization of patients with preconditions on diabetes [8], heart disease [9,10], and hospital readmission for patients with mental or substance use disorders [11]. Recent advances in the machine learning literature have suggested that sparse classifiers, those that use few variables (e.g., 11-regularized Support Vector Machines), have stronger predictive power and generalize better on out-of-sample data points than very complex classifiers [12]. Related work has shown that regularization is equivalent to robustness, that is, learning models which are robust to the presence of outliers in the training set [13]. Moreover, the benefit of using sparse predictors is the enhanced interpretability they provide for both the model and the results.

### 1.1 Objective

Construct data-driven predictive models using data from patients tested for SARS-CoV-2 to predict if a patient will (1) be hospitalized, (2) die, (3) need treatment in an ICU, and/or (4) need a ventilator. To train and test these classifiers we use a public dataset [14] made available by the Mexican government that contains individual information on: demographics (e.g., location), preconditions (e.g., hypertension) and outcomes (e.g., admission to an ICU) for every person who has been tested for SARS-CoV-2 in Mexico.

### 1.2 Main Contributions

- We provide descriptive statistics of the distribution of hospitalized and deceased patients given basic information on preconditions and demographics.
- We develop interpretable models that not only predict the outcomes but also quantify the role of various variables in making these predictions.
- The models we develop leverage data from Mexico. This can motivate additional work using the same data, while the models could be applicable to other Latin American countries with similar population characteristics. This adds to existing work using Electronic Health Records which has focused on patients in the US, Europe, or Asia.

The remainder of the paper is organized as follows: In Section 2 we describe the data used accompanied by descriptive statistics and preprocessing procedures. In Section 3 we describe the binary supervised classification models used and the performance evaluation metrics employed. In Section 3, we present the main results. Discussion of the results can be found in Section 4 and Conclusions in Section 5.

## 2 Data Description and Preprocessing

### 2.1 Data

We use a dataset that has been open for the general public by the Mexican Government (and updated daily) [14]. These data include information about every person who has been tested for SARS-CoV-2 in Mexico. They include demographic information such as: Age, Location, Nationality, the use of an indigenous language; as well as information on pre-existing conditions, including whether the patient has: diabetes, chronic obstructive pulmonary disease (COPD), asthma, immunosuppression, hypertension, obesity, pregnancy, chronic renal failure, other prior diseases, and whether was or is using tobacco. In addition, the data report the dates on which the patient first noticed symptoms, the date when the patient arrived to a care unit, and the date when the patient was deceased (if applicable). Finally, it contains fields showing the result of the SARS-CoV-2 test, weather the patient was hospitalized, has pneumonia, needed a ventilator, and if she/he was treated in an ICU.

As of May 1st, 2020, the data contain more than 91,179 observations out of which more than 20,737 account for positive tests, around 15,000 tests are being processed, and the rest are negative test results. Table 2 1 provides a more precise description of the dataset.

**Table 1:** Descriptive statistics of data set as on May 1st, 2020.

|  |  |
| --- | --- |
| **Total number of tests** | **91,179** |
| Positive | 20,737 |
| Waiting for Result | 15,445 |
| Negative | 54,997 |
| **Total number of patients hospitalized** | **24,099** |
| Positive | 8,221 |
| Waiting for Result | 4,389 |
| Negative | 11,489 |
| Pneumonia | 14,462 |
| Need Ventilator | 1,809 |
| Need ICU | 2,059 |
| **Number of observations with pre-conditions with non-negative test** |  |
| Diabetes | 6,042 |
| COPD | 825 |
| Asthma | 1,235 |
| Immunosuppression | 632 |
| Hypertension | 7,238 |
| Pregnant | 221 |
| Cardiovascular disease | 991 |
| Obesity | 6,998 |
| Chronic renal insufficiency | 820 |
| **Demographics of patients with non-negative test** |  |
| Contact with a positive COVID case | 11,355 |
| Speak an indigenous language | 466 |

### 2.2 Basic Analytics

We provide plots that help us observe trends in the data. We begin by disaggregating data into age groups. In the lower plot of Figure 1 the number of observations of patients having a positive test or waiting their result per age is shown. In addition, the upper bar plot denotes the percentage of the patients in a certain age range who have been hospitalized. This information is aligned with the current knowledge on COVID-19, which indicates that older people have higher risk of being hospitalized. Also, this plot suggests that the risk of being hospitalized increases linearly from the age of thirty up to seventy-five and then plateaus. We ran an ordinary linear regression (OLS) to calculate the rate at which the percentage of hospitalization increases for every additional year of age. The result indicates that the rate is 0.014 with an R^2^ equal to 0.99. This suggests that the risk of hospitalization increases by approximately 1.4% for every year of age between 30 and 75 years old.

**Figure 1:**
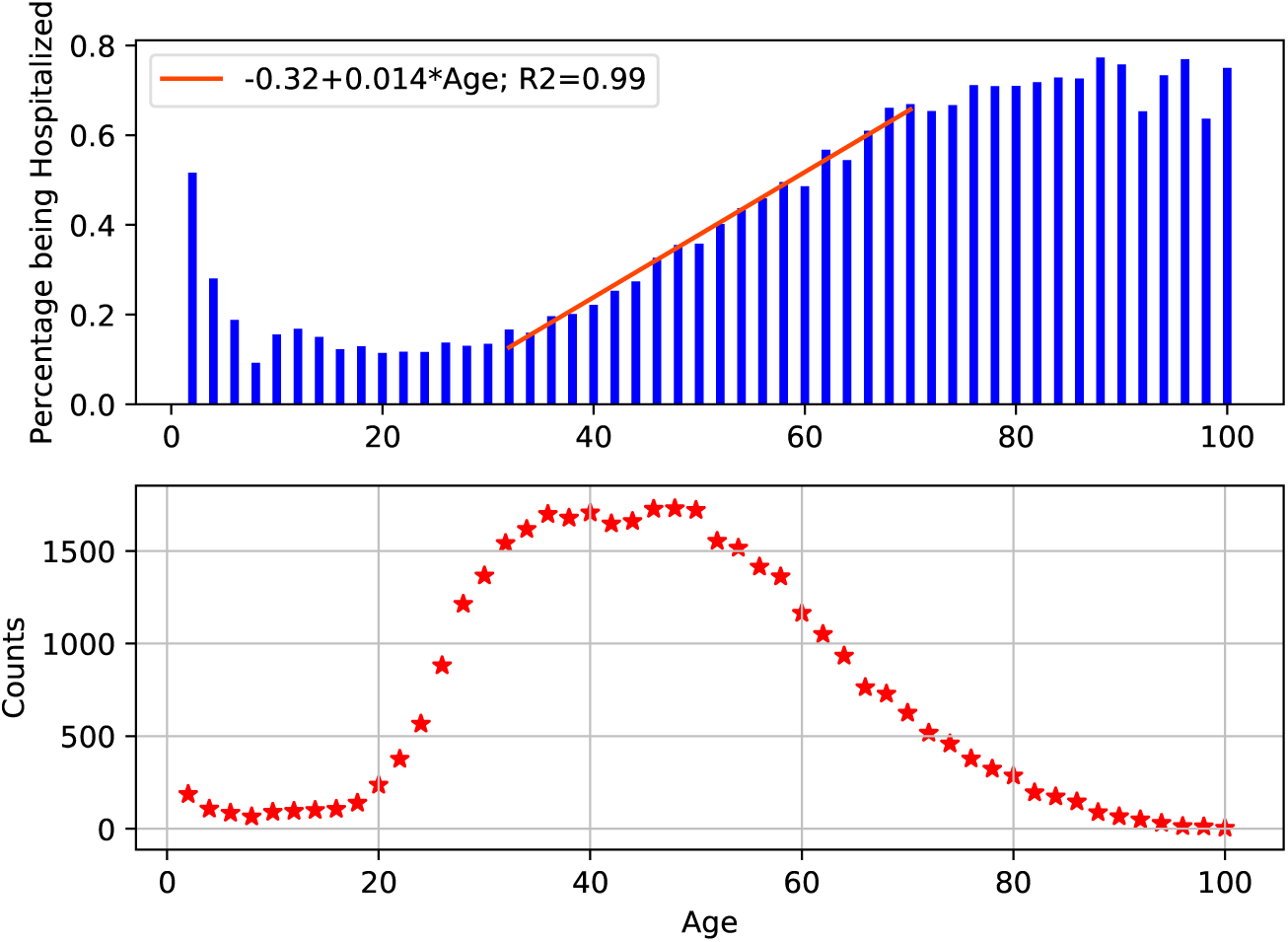
Lower: Number of patients tested positive or waiting for result by age; Upper: Percentage of these patients that have been hospitalized.

Next, in Figure 2 we report the fraction of patients who have been hospitalized, deceased, needed an ICU or a ventilator given a certain precondition. We observe that for both hospitalizations and deaths, preconditions such as chronic renal insufficiency, COPD, diabetes, immunosuppression, cardiovascular disease and hypertension are critical. Nevertheless, even though this gives us information about the risk of a precondition, it does not include the sensitivity regarding how age and preconditions affect a patient with COVID-19.

**Figure 2:**
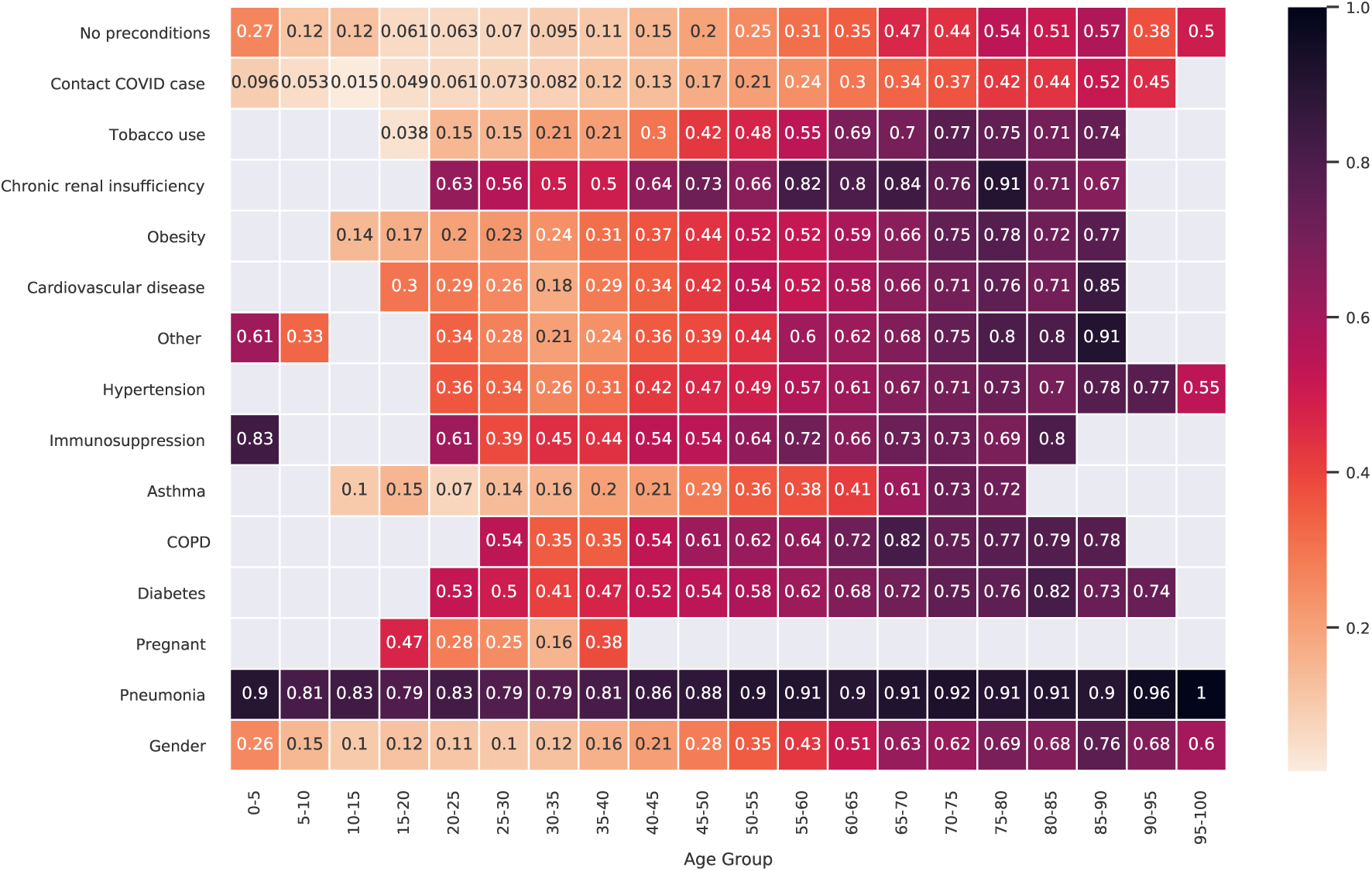
Fraction (%) of patients with a precondition that have been hospitalized, have died or required an ICU or ventilator.

To complement the previous table, we report the same metric by age group and by existing preconditions in Figure 3. To that end, we create age groups for every five years and report results for groups with at least ten observations, otherwise the bin is left blank. On the top row of the table, we include the statistic for a patient without any preconditions. We observe that chronic renal insufficiency, diabetes and immunosuppression are among the preconditions that are associated with a higher hospitalization rate.

**Figure 3:**
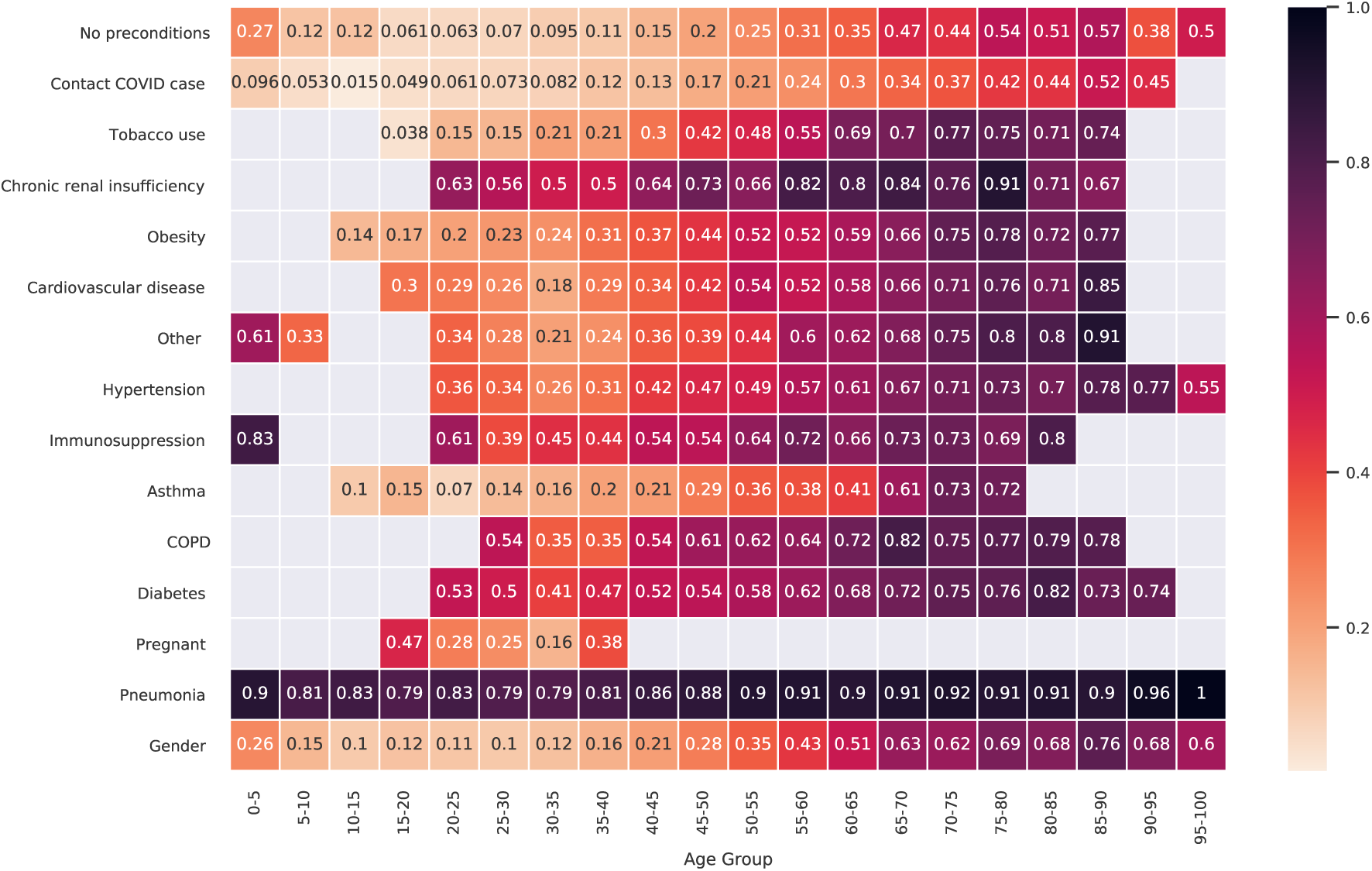
Fraction of population per age being hospitalized given a precondition.

Finally, we present histograms reporting the lag times among various states of the disease for the Mexican population. For this analysis, we separate the data in three groups: individuals with ages between 0-20, 20-50, and patients over 50 years old. In Figure 4 (left), we plot the distribution of the number of days between the onset of symptoms and a subsequent hospitalization. Figure 4 (center) depicts the distribution of time (days) between hospital admission and death. Interestingly, we observe that a large portion of the patients who were hospitalized died the same day they were admitted, potentially suggesting that deterioration of a patient’s condition is abrupt [15,16]. The rest of the distribution behaves like the tail of a Weibull distribution with very few patients being hospitalized for more than three weeks. Finally, Figure 4 (right) shows the distribution of the number of days between the onset of symptoms and death (the mean is 9.8 days).

**Figure 4:**
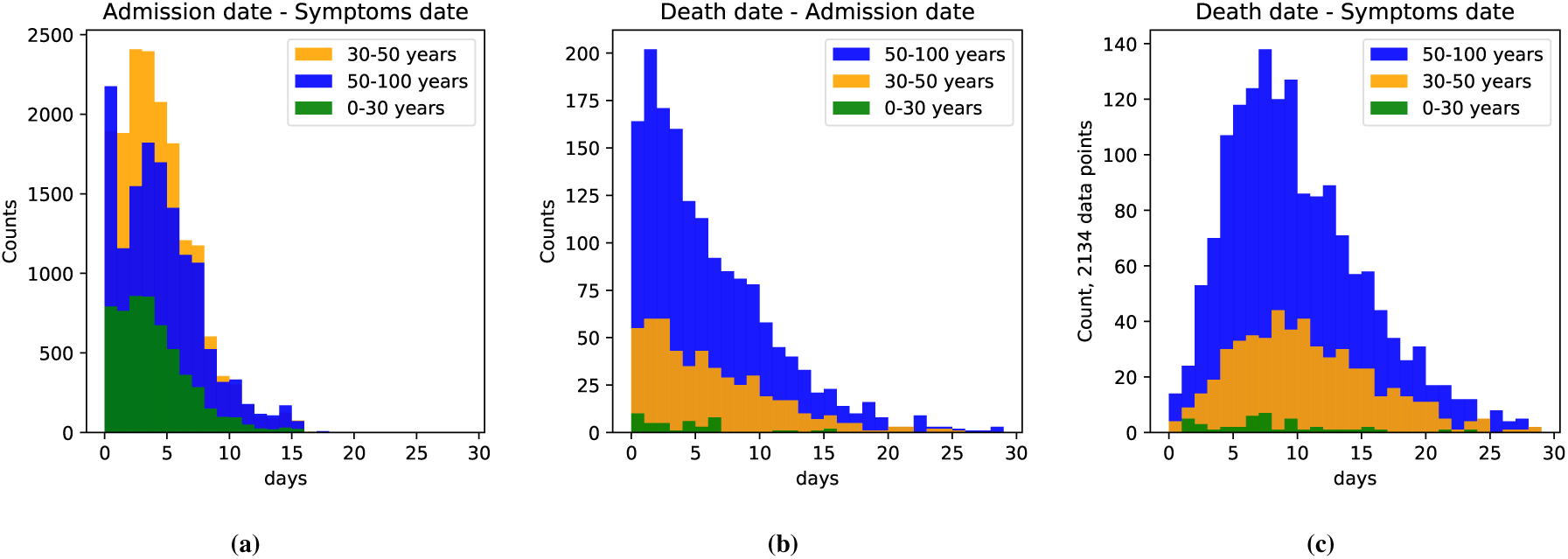
Histograms showing (a) the time between the onset of symptoms and admission date, (b) the time between hospital admission and death, and (c) the time between the onset of symptoms and death.

### 2.3 Preprocessing

#### 2.3.1 Removing outliers

We found a few outliers which are easily identified, for example, the pregnancy of male patients, the date of death of a patient being earlier than the day the patient was admitted to the hospital. Such data points were removed from the dataset.

#### 2.3.2 One-hot encoding

The data contain precondition features reported as categorical. Specifically, each of these precondition features takes the value yes, no, unknown or unspecified. We generate one-hot encoding for all these features. One-hot encoding converts the categorical feature to multiple binary variables by creating auxiliary variables that help distinguish between the different categories of a feature. For the case of our data, one-hot encoding generates three binary variables for each specific precondition; these variables (as opposed to categories) are: no, unknown and unspecified. Then, for each observation, at most one of these variables will be active, pointing to the correct value for the original feature. If none of the three is active, then the value of the precondition is yes.

#### 2.3.3 Removing correlated variables

We find and delete variables that are highly correlated since they, in general, provide similar information. Specifically, we compute pairwise correlations among the variables, and remove one variable from each highly correlated pair (using a threshold of 0.8 for the absolute correlation coefficient). We found that the correlated binary features were the ones corresponding to unknown or unspecified for preconditions. This is because observations that contain an unknown or unspecified value, typically have this same value for all preconditions (not just for one), indicating potential issues in data gathering. Hence, we remove all these auxiliary variables denoting unknown or unspecified preconditions.

## 3 Methods and Metrics

In this section, we briefly introduce the methodologies used to build the binary classifiers. For each model, we train the classifier using four different supervised classification methodologies: sparse Support Vector Machines (SVM), sparse Logistic Regression (LR), Random Forests (RF) and gradient boosted decision trees (XGBoost). For healthcare applications, the first two are preferable due to their interpretability. In turn, the last two are the state-of-the-art classification algorithms today and will serve as a basis to compare the accuracy of the interpretable methods with the non-interpretable benchmark models. Appendix B provides details on these methods, particularly because the robust/sparse LR and SVM formulations are not standard.

### 3.1 Cross-Validated Recursive Feature Elimination

Classifiers based on few variables are desirable because they have stronger predictive power, generalizing better out-of-sample, and offering enhanced interpretability. Aiming to reduce the number of variables, we employ a Recursive Feature Elimination (RFE) procedure [17] to find the variables that optimize a given performance metric. The general framework of this algorithm begins by building a classifier using all the features and computing an importance score for each predictor. In the case of Logistic Regression or Linear SVM, we use as important score the absolute value (or magnitude) of the linear coefficient *β_i_* of feature *i*. After this step, the least important feature (the one with the smallest *\β_i_|*) is deleted from the dataset. We repeat iteratively this process until we are left with one feature. Then, for each of these iterations we report the performance of the model and we pick the set of features that maximize this value. Additionally, at each iteration, we use cross-validation to tune the hyper parameters of the classifier to achieve the best performance.

### 3.2 Performance Evaluation

The primary objective of learning a classifier is to maximize the prediction accuracy, and in our health care setting offer interpretability of the results.

We characterize the prediction accuracy of a classifier using two commonly used metrics: (1) the false positive (or false alarm) rate which measures how many patients were predicted to be in the positive class, e.g., hospitalized, while they truly were not, as a fraction of all negative class patients. In the medical literature, the term specificity is often used and it equals 1 minus the false positive rate. (2) The detection rate that captures how many patients were predicted to be on the positive class while they truly were, as a fraction of all positive class patients. In the medical literature, the detection rate is often referred to as sensitivity or recall. Another term commonly used is precision defined as the ratio of true positives over true and false positives.

A single metric that captures both types of error is the Area Under the Curve (AUC) of the Receiver Operating Characteristic (ROC). ROC plots the detection rate (or sensitivity or recall) over the false positive rate. A naïve random selection (assigning patients to classes randomly) has AUC of 0.5 while a perfect classifier an AUC of 1.

To complement the AUC metric, we report an accuracy metric that computes the ratio of the number of correct predictions over all predictions. Additionally, we compute the F1 score for each class, which is the harmonic mean of precision and recall for that class. We report the weighted F1 score which takes a weighted average of the two per-class F1 scores using as weights the support of each class (normalized over all samples). We finally note that all metrics we report are computed on a randomly selected test set of patients (i.e., out-of-sample) which has not been used for training the models.

## 4 Results

We build binary classification models to predict hospitalization, mortality and the need for an ICU or ventilator. At a minimum, all models use a set of base features composed by: age, gender, diabetes, COPD, asthma, immunosuppression, hypertension, obesity, pregnancy, chronic renal failure, tobacco use, other disease, as well as the SARS-CoV-2 test result which is either positive or pending (we exclude all negative cases to train our models). In this section, we provide a summary of the results while in the Appendix A we provide all results.

### 4.1 Hospitalizations

Our first model predicts if a patient who has tested positive or is waiting for the test result will be hospitalized given their base features. This model has a moderate accuracy for all methodologies employed which accounts for an AUC of 0.62 and an accuracy of classifying 61The coefficients of the SVM and LR models have the same trend and suggest that the features that contribute the most for predicting the hospitalization of a patient are: age, gender, chronic renal insufficiency, diabetes, immunosuppression or if the patient is pregnant. The rest of the variables (COPD, Obesity, Hypertension, Other, Tobacco Use, Cardiovascular disease and Asthma) have a much smaller impact. It is however possible that some of these variables have smaller coefficients because the effect is captured by another highly correlated variable (e.g., obesity and diabetes).

### 4.2 Mortality

We explore two models to predict mortality. The first model assumes we only know the base features of a patient whereas the second model includes variables that indicate if the patient has been hospitalized or not, has pneumonia, or has needed an ICU or ventilator. The reason to consider the first model is to have a classifier which identifies which patients are the most vulnerable prior to hospitalization, while the second model predicts the mortality of an individual in the hospital by using information on how the disease is progressing. In order to have a more balanced dataset and to detect better the deceased class, we ran this model only on the observations of patients who have been hospitalized and have been tested positive or are waiting for their test result.

**Prior to attending a healthcare facility**. This model considers the case in which we only know the base features of a patient. When running this model, we are able to predict with 73% accuracy and with an AUC equal to 0.69 the mortality of a patient.

**After attending a healthcare facility**. We also consider the case in which we have information about the hospitalization, pneumonia ICU and ventilator of a patient. This classification task achieves an AUC of 0.74 with an accuracy of 76%.

Both interpretable models, LR and SVM, suggest that the variables that are critical for predicting mortality are the patient’s age, test status, immunosuppression and pregnancy. For the model that has more features, as expected, information about the need for ventilator and ICU are highly relevant when predicting mortality.

### 4.3 ICU need

Similar to the mortality case, we train two classification models to predict the need for an ICU.

**Prior to knowing if patient has developed pneumonia**. By only using the base features, we achieve an accuracy of 80% with an AUC of 0.54.

**Knowing if a patient has developed pneumonia or not**. The results for this model suggest that information about pneumonia is relevant for predicting ICU need as it raises the accuracy of the model to 82% and the AUC to 0.63.

In these cases, SVM and LR suggest that information on: development of pneumonia (if available), cardiovascular disease, asthma, and test result are among the features with higher importance for predicting the need for an ICU.

### 4.4 Ventilator Need

Similar to the mortality and ICU models, we develop two versions of the model.

**Prior to knowing if patient has developed pneumonia or needs an ICU**. The accuracy of this model is higher than both the mortality and the ICU models, achieving an accuracy of 81% and an AUC of 0.56.

**Knowing if a patient has developed pneumonia or not and the need for an ICU**. This model suggests, as expected, that this additional information is relevant for predicting ventilation need. It increases its accuracy to 83% and the AUC to 0.77.

As in the mortality case and the ICU case, both interpretable models are consistent and have an accuracy comparable or higher than RF and XGBoost. Moreover, both models classifying the need for a ventilator show that information on ICU and pneumonia (if available), age, gender, cardiovascular disease, obesity, pregnancy, and test result are the most relevant features for predicting the need for a ventilator given that a patient has tested positive or is waiting for a test result.

## 5 Discussion

Overall, the models we develop range from moderately to significantly accurate. Predicting hospitalizations appears harder just based on the basic variables at our disposal, particularly considering all patients who have a positive test or with a test pending. Potential additional features are at play including state of health (measured through detailed lab results) and the viral load they were exposed to. Furthermore, a number of hospitalizations are driven by socioeconomic factors, e.g., the living arrangements of a patient and whether he/she can pose infection risk for many others. Still, an AUC of 0.62 is significantly better than random and the results could help tighten estimates on the number of hospitalizations expected.

From an actionable and planning perspective, predicting ICU treatment and ventilator need are quite useful. These models can be quite accurate, achieving accuracies of 82% and 84%, respectively, when information on how the disease is progressing is taken into account (e.g., development of pneumonia). Similarly, the mortality model can achieve an accuracy of 76%. Again, it is important to emphasize that we lack very important information, such as lab results, which can characterize the state of the patient prior to hospitalization and throughout its duration.

An interesting observation is that interpretable models (such as LR and SVM), when used in conjunction with robustness/regularization approaches and elaborate feature selection procedures, can lead to performance that is comparable, if not better than more complex and expensive classifiers. The significant advantage of the former models is that they are interpretable and provide information on which variables drive the predictions.

To the extent that these risk models can be used to prioritize the use of resources, we understand that medical risk is not the only factor in making such decisions. Nevertheless, in order to quantify medical risk one can leverage the models presented in this work.

## 6 Conclusion

We develop models to identify the medical risk of a patient with (or suspected for) COVID-19. We hope this work can help hospitals and policymakers to distribute more effectively their limited resources including tests, ICU beds and ventilators, as well as, to motivate countries and healthcare systems to standardize and share data with the medical informatics community. Moreover, we hope this research spreads the knowledge of the existence of this public dataset and motivates researchers to work with these data. Finally, we hope that risk models are taken into account to fine-tune social distancing advisories, moving from “blanket” to risk-based, as well as prioritizing vaccine distribution to the more vulnerable and to those who need to interact with the more vulnerable. For the sake of reproducibility and to facilitate the analysis for further research we have made open source our models and results on a Github repository [18].

## Data Availability

Data from a publicly available repository, updated daily, containing information from approximately 91,000 patients in Mexico were used. The data for each patient include demographics, prior medical conditions, SARS-CoV-2 test results, hospitalization, mortality and whether a patient has developed pneumonia or not.

https://www.gob.mx/salud/documentos/datos-abiertos-152127

## 7 Acknowledgements

Research partially supported by the NSF under grants IIS-1914792, DMS-1664644, and CNS-1645681, by the ONR under MURI grant N00014-19-1-2571, and by the NIH under grant 1R01GM135930.

The authors would like to thank Diana Sverdlin-Lisker at the Massachusetts Institute of Technology for her useful discussions and for proofreading this work.

### 9 Appendix A

#### 9.1 Hospitalizations

|  | SVM | LR | RF | XGBoost |
| --- | --- | --- | --- | --- |
| Accuracy | 0.609 | 0.609 | 0.591 | 0.606 |
| F1w | 0.607 | 0.607 | 0.593 | 0.601 |
| AUC | 0.622 | 0.622 | 0.612 | 0.620 |

Coefficients:
|  | SVM | LR |
| --- | --- | --- |
| Age | 1.000 | 1.000 |
| Pregnant | 0.166 | 0.172 |
| Chronic Renal Insufficiency | 0.156 | 0.167 |
| Diabetes | 0.181 | 0.165 |
| Immunosuppression | 0.139 | 0.139 |
| COPD | 0.097 | 0.094 |
| Obesity | 0.081 | 0.083 |
| Other | 0.046 | 0.046 |
| Hypertension | 0.045 | 0.039 |
| Tobacco Use | 0.007 | 0.007 |
| Cardiovascular Disease | -0.008 | -0.005 |
| Asthma | -0.062 | -0.065 |
| Gender | -0.119 | -0.121 |

#### 9.2 Mortality

##### 9.2.1 Prior to attending a healthcare facility

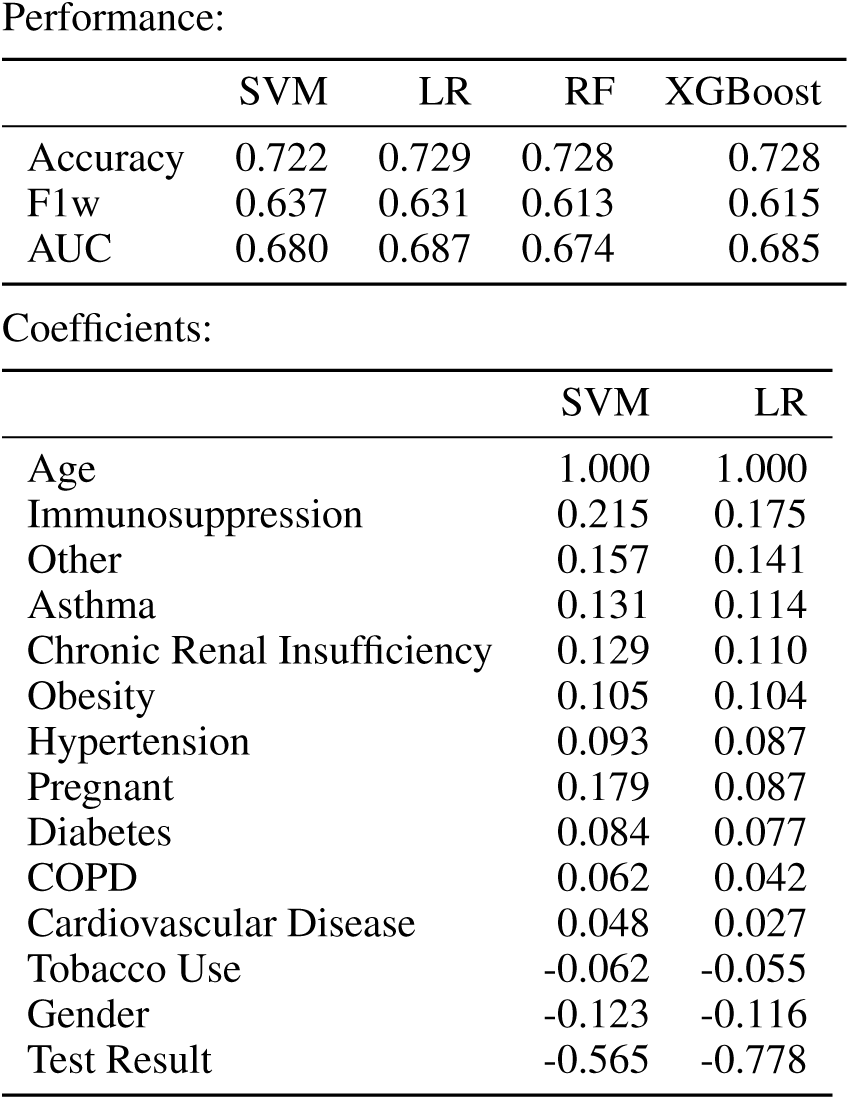

##### 9.2.2 After attending a healthcare facility

9.2.2 After attending a healthcare facility Performance:
|  | SVM | LR | RF | XGBoost |
| --- | --- | --- | --- | --- |
| Accuracy | 0.761 | 0.762 | 0.752 | 0.762 |
| F1w | 0.717 | 0.711 | 0.647 | 0.705 |
| AUC | 0.729 | 0.744 | 0.744 | 0.752 |

Coefficients:
|  | SVM | LR |
| --- | --- | --- |
| Age | 1.000 | 1.000 |
| Ventilator | 0.638 | 0.486 |
| Pregnant | 0.294 | 0.222 |
| Immunosuppression | 0.222 | 0.200 |
| Other | 0.147 | 0.139 |
| Asthma | 0.146 | 0.139 |
| Obesity | 0.136 | 0.132 |
| Chronic Renal Insufficiency | 0.122 | 0.112 |
| Pneumonia | 0.103 | 0.108 |
| Hypertension | 0.089 | 0.086 |
| Diabetes | 0.090 | 0.086 |
| ICU | 0.095 | 0.067 |
| COPD | 0.067 | 0.051 |
| Cardiovascular Disease | -0.011 | -0.012 |
| Tobacco Use | -0.077 | -0.069 |
| Gender | -0.096 | -0.095 |
| Test Result | -0.545 | -0.718 |

#### 9.3 ICU Need

##### 9.3.1 Prior to knowing if patient has developed pneumonia

|  | SVM | LR | RF | XGBoost |
| --- | --- | --- | --- | --- |
| Accuracy | 0.799 | 0.799 | 0.799 | 0.799 |
| F1w | 0.710 | 0.710 | 0.710 | 0.710 |
| AUC | 0.538 | 0.548 | 0.541 | 0.554 |

Coefficients:
|  | SVM | LR |
| --- | --- | --- |
| Age | 1.000 | 1.000 |
| Obesity | 0.532 | 0.502 |
| Pregnant | 0.830 | 0.488 |
| Cardiovascular Disease | 0.564 | 0.455 |
| Asthma | 0.457 | 0.364 |
| Other | 0.223 | 0.170 |
| Diabetes | 0.064 | 0.052 |
| Hypertension | -0.000 | 0.000 |
| Immunosuppression | -0.202 | -0.159 |
| COPD | -0.277 | -0.209 |
| Tobacco Use | -0.266 | -0.250 |
| Chronic Renal Insufficiency | -0.340 | -0.316 |
| Gender | -0.372 | -0.368 |
| Test Result | -0.777 | -0.843 |

##### 9.3.2 Knowing if a patient has developed pneumonia

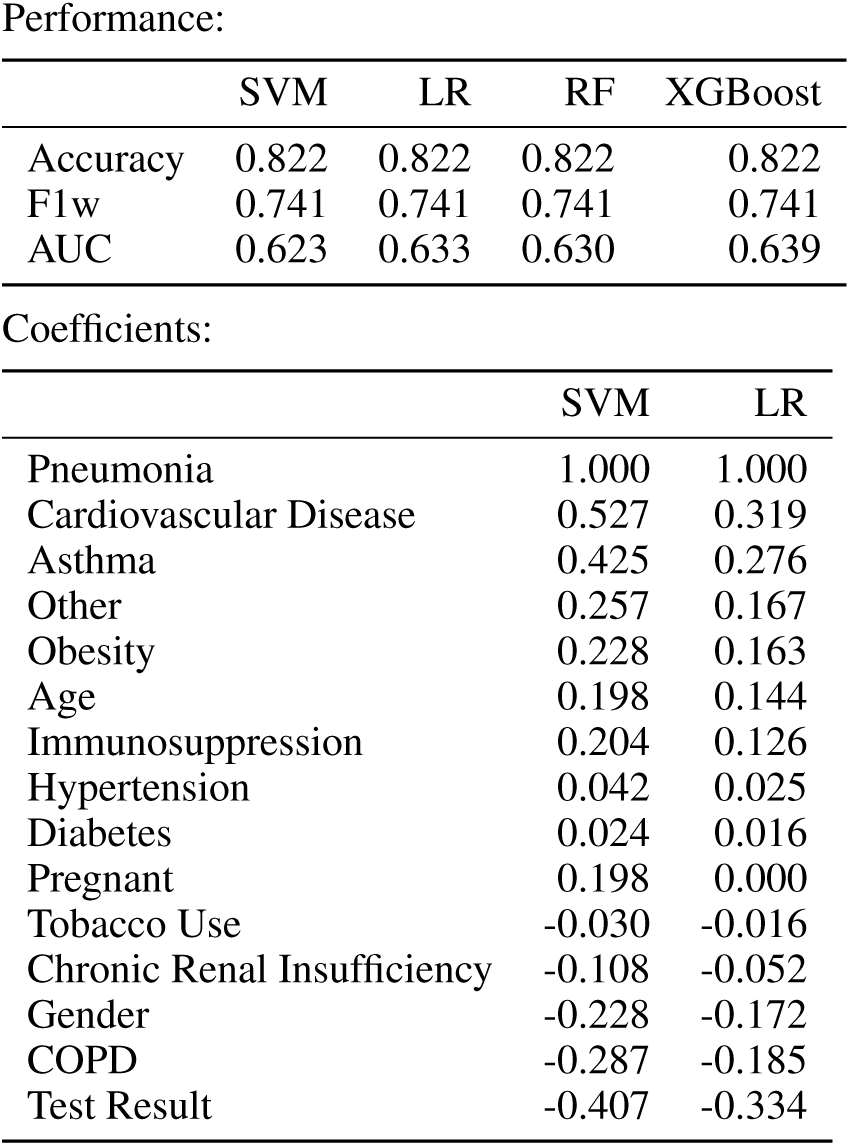

#### 9.4 Ventilator Need

##### 9.4.1 Prior to knowing if patient has developed pneumonia or needs an ICU

|  | SVM | LR | RF | XGBoost |
| --- | --- | --- | --- | --- |
| Accuracy | 0.805 | 0.805 | 0.805 | 0.805 |
| F1w | 0.718 | 0.718 | 0.718 | 0.718 |
| AUC | 0.557 | 0.560 | 0.541 | 0.560 |

Coefficients:
|  | SVM | LR |
| --- | --- | --- |
| Age | 1.000 | 1.000 |
| Obesity | 0.595 | 0.519 |
| Cardiovascular Disease | 0.360 | 0.254 |
| Tobacco Use | 0.135 | 0.097 |
| Hypertension | 0.090 | 0.081 |
| Diabetes | -0.009 | 0.000 |
| Chronic Renal Insufficiency | -0.018 | 0.000 |
| COPD | -0.063 | -0.017 |
| Immunosuppression | -0.162 | -0.134 |
| Other | -0.198 | -0.184 |
| Asthma | -0.261 | -0.196 |
| Pregnant | -0.486 | -0.297 |
| Gender | -0.396 | -0.387 |
| Test Result | -0.748 | -0.769 |

##### 9.4.2 Knowing if a patient has developed pneumonia or not and the need for an ICU

|  | SVM | LR | RF | XGBoost |
| --- | --- | --- | --- | --- |
| Accuracy | 0.827 | 0.830 | 0.818 | 0.825 |
| F1w | 0.810 | 0.809 | 0.736 | 0.791 |
| AUC | 0.774 | 0.773 | 0.770 | 0.779 |

Coefficients:
|  | SVM | LR |
| --- | --- | --- |
| ICU | 1.000 | 1.000 |
| Pneumonia | 0.233 | 0.724 |
| Age | 0.174 | 0.275 |
| Chronic Renal Insufficiency | 0.072 | 0.122 |
| Obesity | 0.066 | 0.113 |
| Cardiovascular Disease | 0.064 | 0.097 |
| Tobacco Use | 0.029 | 0.043 |
| Pregnant | -0.079 | 0.000 |
| Hypertension | -0.012 | -0.013 |
| COPD | -0.013 | -0.013 |
| Diabetes | -0.012 | -0.020 |
| Immunosuppression | -0.056 | -0.034 |
| Other | -0.047 | -0.074 |
| Gender | -0.045 | -0.081 |
| Asthma | -0.061 | -0.083 |
| Test Result | -0.064 | -0.117 |

### 10 Appendix B

For all models we will assume we are given training data x*_i_* ∊ *R^D^*, where we use bold letters to denote vectors, and classification labels *y_i_* = {0,1} for all *i =* 1,…, *n*, where *D* is the number of variables in the data set, x*_i_* is the vector of variables for the *i*-th patient, and n is the number of samples (or patients).

#### 10.1 Sparse Linear Support Vector Machines

A support vector machine (SVM) is a binary classifier that seeks to find a separating hyperplane in the feature space, so that the two classes reside on opposites sides [24]. The main idea of the SVM is to maximize the margin between the data and the chosen hyperplane, where the margin is defined as the distance of the closest data point in a class to the margin. Unfortunately, in many cases the data are not linearly separable, meaning that there is no hyperplane able to perfectly separate all points. The so-called soft-margin SVM tolerates this misclassification, and it is formulated as follows:

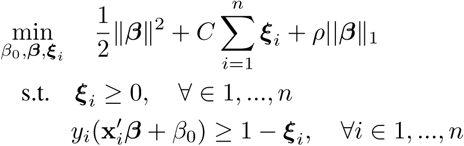

where the hyperplane is characterized by the perpendicular vector and the intercept (*β*, *β*_0_), and the variables are used to identify the misclassification of a point which is penalized by C. For SVM, the labels are assumed to be in {−1, 1}. What is different than a standard SVM formulation is the use of an 11-norm regularizer, inspired by robustness arguments [13]. The scalar ρ represents the strength of the regularizer. This problem can be reformulated as a convex quadratic programming problem which can be solved using standard solvers.

#### 10.2 Sparse Logistic regression

Similar to sparse SVM, logistic regression (LR) [25] is an interpretable binary linear classifier. The key idea is to model the posterior probability of the outcome *y_i_* (e.g. a patient being hospitalized) as a logistic function of a linear combination of the features x*_i_*. To that end, it uses parameters *θ* that weigh the input features and an offset *θ*_0_. These parameters are selected by maximizing the log-likelihood of a function by the use of gradient-based algorithms. LR has been particularly popular in the medical literature because it predicts the probability that a sample belongs to the positive (or negative) class. In this work, we employ a sparse logistic regression. This is an *l*_1_-regularized logistic regression which includes in the objective function an extra term proportional to ||*θ||*_1_ in the log likelihood sense. Same as in sparse SVM, the motivation to include this term is to render the predictor more robust [26]. Formally, the problem is defined as

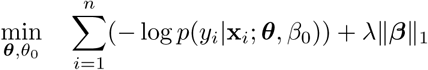

where 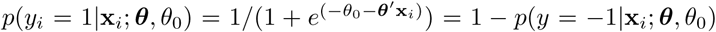, and *λ* is a parameter controlling the sparsity term. When *λ* = 0, we have the standard logistic regression model.

#### 10.3 Random Forests

This type of classifiers is one of the most precise models for binary classification today. Random Forest (RF) [27] are part of a bigger class of predictors called ensemble methods. The main idea of ensemble classifiers is to reduce the variance of an estimated predictor by training many noisy but approximately unbiased models and making the classification decision based on the majority of vote of these weak classifiers. In particular, RF is an ensemble of decision trees (DT) [28]. To grow each DT of the RF, the model uses data obtained through random sampling with replacement from the training set. A DT is fully grown until a minimum size (or depth) is reached. Even if the decision of an individual DT is very noisy due to the sampling process, the average of many DTs is not, as long as these trees are not highly correlated. RFs are easy and fast to implement to large data sets and do not have a high risk of overfitting. A general disadvantage of these methods is the lack of interpretability as every prediction is obtained by majority voting of many (hundreds or thousands) of DTs. weak and small DTs.

#### 10.4 XGBoost

Similar to RF, XGBoost (which stands for Extreme Gradient Boosting) [29] is a decision-tree-based ensemble model that uses a gradient boosting framework. In prediction problems involving unstructured data (images, text, etc.) deep neural networks tend to outperform all other algorithms or frameworks. However, when it comes to small-to-medium structured tabular data, as is the case for our application in hand, decision tree based algorithms are considered best-in-class.

Similar to RF, XGBoost has great accuracy but lacks interpretability and it is included in this work as a benchmark. This classifier, as of today, has been credited with winning numerous Kaggle competitions [30] and has being used widely in cutting-edge industry applications.

